# User Experience Evaluation of Cogscreen for Screening Mild Cognitive Impairment: Formative and Summative Evaluation

**DOI:** 10.1101/2025.10.22.25338470

**Authors:** Eunji Hwang, Hyunsun Ham, Dasom Lee, Hanna Kim, Sang Hoon Oh, Jun Gyu Kang, Sehee Shim, Somin Lee, Jung-Hae Youn, Jun-Young Lee

## Abstract

**Background:** Mild Cognitive Impairment (MCI) is an early stage of cognitive decline and a key risk factor for dementia. Early detection is essential for timely intervention and prevention, so various digital screening tools have been developed recently. However, many of these tools have limitations. Without assistance, users may have difficulty completing the tests, which could result in increased failure rates and reduced accuracy. This study evaluates the usability of Cogscreen, a mobile cognitive screening tool that can be completed independently from start to finish.

**Methods:** A two-step usability evaluation was conducted: a formative evaluation with five physiatrists assessing design clarity and usability issues, and a summative evaluation with 15 participants analyzing task completion rates and System Usability Scale (SUS) scores.

**Results:** The formative evaluation identified strengths in interface design and overall usability, while also noting areas for refinement. The summative evaluation showed a 97% task completion rate and an average SUS score of 80.2, indicating strong usability. Minor issues were reported in personal information entry and graphical result interpretation.

**Conclusions:** Cogscreen demonstrated high usability as a digital screening tool for MCI, underscoring its promise for supporting early detection of cognitive decline. Future refinements should focus on enhancing error handling and improving demographic guidance. Expanding testing across diverse populations could further validate its potential clinical utility.

## Introduction

Mild Cognitive Impairment (MCI), an early stage of cognitive decline, is a key risk factor for dementia, including Alzheimer’s disease (AD).^1,2^ Affecting 10–20% of individuals aged 65 and older, MCI highlights the need for early detection and intervention to delay progression, improve quality of life, and reduce healthcare costs.^3,4^ Recent advances in disease-modifying therapies for Alzheimer’s disease, such as monoclonal antibodies targeting amyloid, have shown that treatment efficacy is greatest in the prodromal or early stages of the disease.^5,6^ Therefore, identifying individuals with MCI at an early stage is crucial to enable timely intervention and maximize the potential benefits of emerging therapeutic approaches.

Although tools such as the Mini-Mental State Examination (MMSE) and the Montreal Cognitive Assessment (MoCA) remain the most widely employed cognitive screening measures, their reliance on trained professionals, time-consuming administration, and limited sensitivity to subtle cognitive decline significantly constrain their utility in early detection.^7,8^

These limitations have driven the development of digital cognitive screening tools that aim to improve accessibility, scalability, and early detection of MCI. The Cambridge Neuropsychological Test Automated Battery (CANTAB) offers culture-independent assessments using non-verbal stimuli.^9^ The Cogstate Brief Battery (CBB) enables repeated cognitive testing with high test-retest reliability.^10^ The Digital Brain Function Screen (DBFS) demonstrates a strong correlation with traditional tools while offering automated administration.^11^

Digital cognitive screening tools offer a promising approach to the early detection of cognitive impairments, but they face limitations in terms of accuracy and usability, particularly when users complete assessments independently. For Older adults, difficulties with comprehension, technological interaction, and task format can compromise both feasibility and validity, often resulting in incomplete or failed assessments and highlighting the need for rigorous usability evaluations in this population. In a study on the MoCA XpressO, 25% of participants failed to complete the test due to comprehension issues and technical difficulties.^12^ Similarly, Feedback from users of the MyCog system revealed frustration with unclear progress tracking and design flaws, further hindering usability and completion rates.^13^ Recurring challenges, such as incomplete task performance in unsupervised settings, limited acceptance among individuals with lower digital literacy, and narrow single domain task coverage, remain unresolved. Positioning Cogscreen in this context, we aim to examine whether a brief, multidimensional, self-administered design can mitigate these barriers.

Recent systematic and scoping reviews have also summarized the broader state of digital cognitive assessment tools for older adults.^14,15^ These reviews highlight several recurring limitation s, including the limited evaluation of individuals with diverse educational and cultural backgrounds, variable psychometric validation across tools, and the rarity of usability testing in real-world or unsupervised settings. Research in primary care settings indicates that although home-based and tablet-based assessments show promise, individuals with limited digital literacy are more likely to encounter difficulties in completing them.^16^ Given the design goals of Cogscreen, this study evaluates whether its design mitigates some of the recurring usability barriers.

Cogscreen was developed in recognition of these challenges to examine whether a brief, multidimensional, self-administered design can mitigate them. Unlike many tools that focus primarily on cognitive tasks alone, Cogscreen combines objective tests with self-report scales for subjective cognitive decline and depressive symptoms, offering a multidimensional perspective. It integrates objective cognitive tasks with self-report scales of subjective cognitive decline (SCD) and depressive symptoms while prioritizing usability and accessibility.^17,18^

It is designed to enable fully independent completion through specific usability features. Unlike traditional paper-and-pencil tests that require trained examiners, Cogscreen was designed for independent administration with a streamlined user interface (UI) and user experience (UX), which minimizes the need for in-person assistance.^19–21^ As usability highly influences the feasibility of digital cognitive assessments, it is essential to evaluate whether individuals can effectively navigate and complete the test as intended.^22^

In this study, we assessed the usability of Cogscreen through both formative and summative evaluations. The formative evaluation aimed to identify usability issues and potential hazards through expert feedback, while the summative evaluation assessed user performance, task completion, and overall usability in a real-world setting.^23–26^

## Method

### Study Design

This study adopted a two-phase evaluation framework, comprising formative evaluation and summative evaluation, designed to systematically assess the usability of Cogscreen (Fig 1). Formative evaluation entails expert-driven analyses conducted during early development to identify design issues and guide iterative improvements, while summative evaluation refers to structured user testing intended to validate the overall usability and safety of the system prior to deployment. The primary objective was to evaluate whether users could operate the tool independently, without assistance, while ensuring the clarity of instructions provided within the application and accompanying materials.

**Figure 1.**
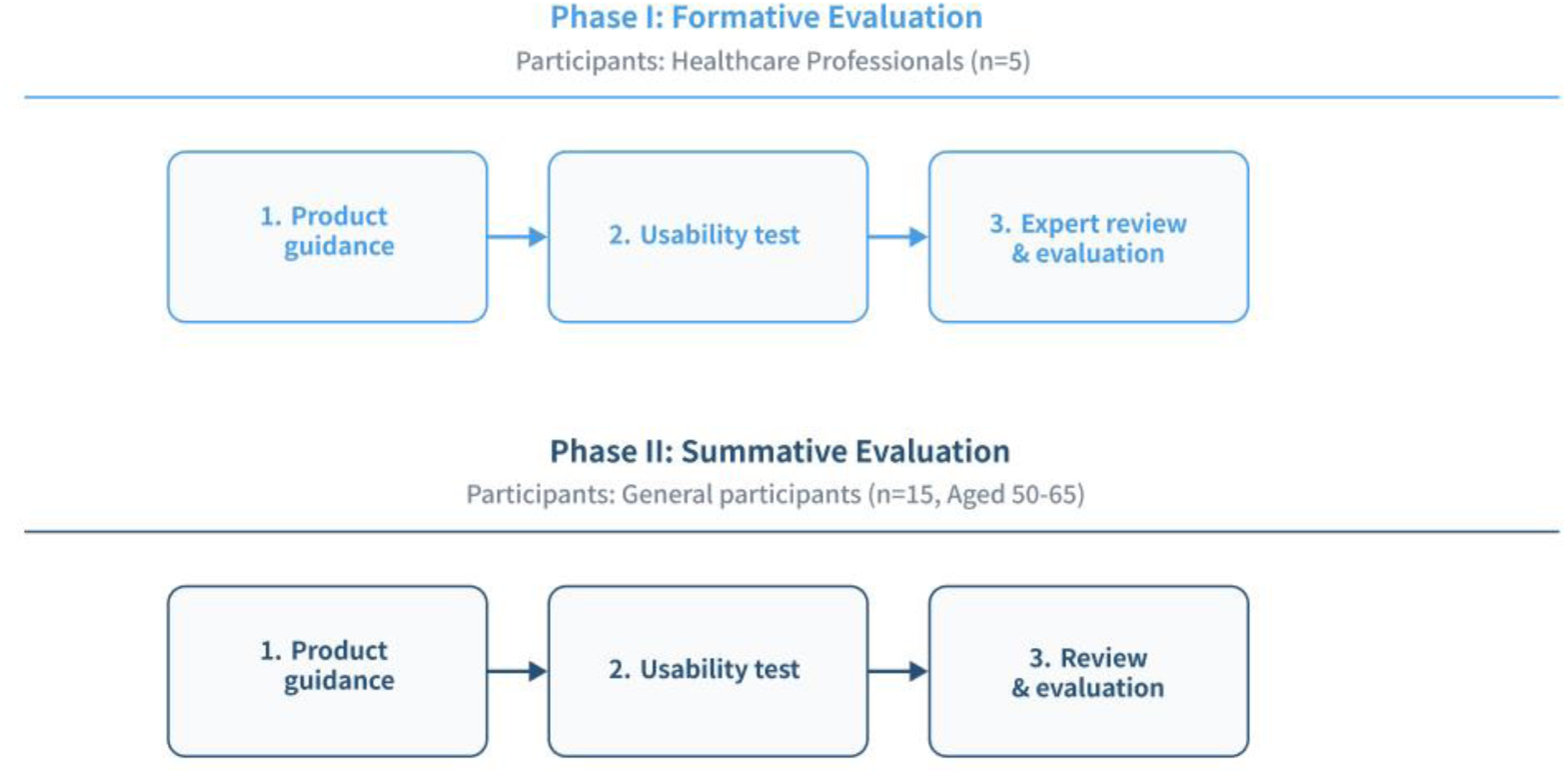
The systematic workflow of Formative and Summative Evaluations

The formative evaluation focused on identifying design issues and several factors for improvement to optimize user experience, while the summative evaluation aimed to validate the tool’s usability by assessing its overall effectiveness, efficiency, and user satisfaction in a real-world setting. Both evaluations adhered to the IEC 62366-1:2015+AMD1:2020, a standard on the application of usability engineering for medical devices.^27^

The study protocol followed a standardized procedure across both evaluation phases. Participants were provided with a user manual of Cogscreen and were informed about potential risks prior to completing Cogscreen and participating in post-evaluation interviews. User interactions with the tool were observed through a one-way mirror, and usage patterns were systematically recorded for analysis.

### Tool Description

Cogscreen is a mobile cognitive screening tool developed to facilitate independent assessment of cognitive function without reliance on trained personnel. The tool is accessible on personal mobile devices and consists of a series of tasks requiring approximately five to ten minutes to complete. Upon completion, users receive detailed feedback on their cognitive performance along with personalized recommendations based on task outcomes.

Cogscreen includes two cognitive tests designed to assess key domains of cognitive functioning. The first task, a cued word recall test, evaluates memory retention by measuring the user’s ability to recall recently presented information. The second task, a digit-symbol substitution test, examines executive function and processing speed by requiring users to match symbols to their corresponding digits under time constraints. In addition to these tests, Cogscreen incorporates a self-report questionnaire that captures users’ subjective cognitive concerns and depression-related factors, providing valuable context for interpreting task performance. The detailed task flow and user interface were presented in Korean. English-translated versions are available upon request from the corresponding author.

### Formative Evaluation

The formative evaluation was conducted with five physiatrists from Wonju Severance Christian Hospital Medical Device Clinical Trial Center (Table 1). All expert participants also provided written informed consent before participating in the formative evaluation. After providing the participants with the product manual and usage training, the product was evaluated by completing evaluation sheets. These sheets used a 5-point Likert scale, with higher scores reflecting more favorable usability assessments. This formative evaluation aimed to assess the suitability of the design for intended users, identify factors that facilitate or hinder its use, and gather expert feedback on barriers to guide subsequent design refinements. In addition to completing structured evaluation sheets, the five physiatrists provided qualitative feedback on clinical applicability, safety considerations, and potential barriers for patients with cognitive impairment. Their input focused on whether the instructions, interface design, and error management functions would be understandable and practical in real-world clinical contexts.

**Table 1.** Demographic information in formative and summative evaluations.

|  | Healthcare Professionals (n=5)<br>in formative evaluation | General Participants (n=15) in<br>summative evaluation |
| --- | --- | --- |
| **Age (%)** |  |  |
| 20s | 20% (n=1) | 0% (n=0) |
| 30s | 40% (n=2) | 0% (n=0) |
| 40s | 20% (n=1) | 0% (n=0) |
| 50s | 20% (n=1) | 20% (n=3) |
| 60s | 0% (n=0) | 80% (n=12) |
| **Gender (%)** |  |  |
| Male | 80% (n=4) | 40% (n=6) |
| Female | 20% (n=1) | 60% (n=9) |
| **Occupation (%)** |  |  |
| Physiatrists | 100% (n=5) | n/a |
| Homemaker | n/a | 33.3% (n=5) |
| Self-employed | n/a | 26.7% (n=4) |
| Office worker | n/a | 13.3% (n=2) |
| Unemployed | n/a | 20% (n=3) |
| Caregiver | n/a | 6.7% (n=1) |

| Similar Product Experience (%) |  |  |
| --- | --- | --- |
| Yes | 60% (n=3) | 0% (n=0) |
| No | 40% (n=2) | 100% (n=15) |

In a simulated use environment, the trainer provided education on the device to healthcare professionals. Afterward, the healthcare professionals used the device themselves to analyze the product and collected expert opinions using the provided evaluation forms. One facilitator, one observer, and five subjects who understand the principle of the device to be evaluated participated. The facilitator performed procedures such as explaining the purpose of the test, confirming participation, explaining the need for confidentiality, explaining the risks and protections related to the test, and post-interview according to the protocol. An observer recorder observed and recorded participants’ behavior, and used errors and opinions outside the testing room equipped with a one-way mirror.

### Summative Evaluation

Feedback was provided during the formative evaluation. Most comments focused on optimizing user experience rather than addressing critical usability or safety concerns. Relevant insights were considered during the preparation of the summative evaluation, which was conducted to assess the tool’s usability in a real-world setting.

The summative evaluation involved fifteen participants aged 50 to 65, recruited through Wonju Severance Christian Hospital Medical Device Clinical Trial Center (Table 1). The sample size (n = 15) was determined based on established usability study guidelines. Although IEC 62366 1 does not specify a required sample size, both the FDA and human factors research recommend approximately 15 participants for summative usability evaluations of medical or cognitive interface systems to identify most usability issues.^28^ Therefore, this number was considered sufficient for an exploratory usability evaluation. Participants were screened based on exclusion criteria, which ruled out individuals with hearing or visual impairments, illiteracy, diagnosed brain disorders, cerebral palsy, neurological or physical conditions affecting cognition, acute or chronic infections, or severe mental illnesses.

In Table 1, ‘similar product experience’ was defined as any prior use of digital cognitive assessment tools or mobile applications designed for memory, attention, or other cognitive tasks. This variable applied to both expert and lay participants and was assessed through self-report during participant screening. All participants were cognitively normal older adults without a diagnosis of mild cognitive impairment or dementia. This study was therefore designed as a first step to evaluate the usability of Cogscreen in a non-clinical older adult population before extending to individuals with cognitive disorders.

All post-evaluation interviews followed a semi-structured format, using a predefined set of open-ended questions that explored participants’ overall experiences with Cogscreen, perceived usability, and suggestions for improvement. During the scenario-based tasks, participants were encouraged to provide subjective opinions and recommendations both during and after each scenario. Following each task, they were invited to comment on the clarity of the user manual, convenience or challenges experienced while using the system, and desired additional features. Participants were also encouraged to share any further feedback in an open discussion. Interview data were analyzed using qualitative content analysis, with two independent researchers coding the data to identify key patterns and themes related to usability, accessibility, and user satisfaction. Detailed interview questions and sample responses are provided in Supplementary Table 2. Before the evaluation, the study coordinator explained the purpose of the test, obtained written informed consent, described potential risks and protective measures, and assured participants of confidentiality. Participants were provided with a general overview of the product and completed assigned tasks independently. After completing the tasks, one-on-one interviews were conducted to collect detailed feedback, and the System Usability Scale (SUS) was used to assess user satisfaction.

## Results

### Formative Evaluation

The formative evaluation systematically examined documentation, usability, and satisfaction, providing a structured overview of both strengths and areas requiring refinement (Table 2). The Documentation and Clarity of User Guidance highlighted strong performance in defining intended purposes with a mean score of 4.8 ± 0.4, clarity of manuals with 4.8 ± 0.4, and usage methods with 4.8 ± 0.4. However, defining potential users scored lower at 4.0 ± 1.8, and guidance on educational levels for users was identified as an area needing improvement at 3.8 ± 1.6.

**Table 2.**
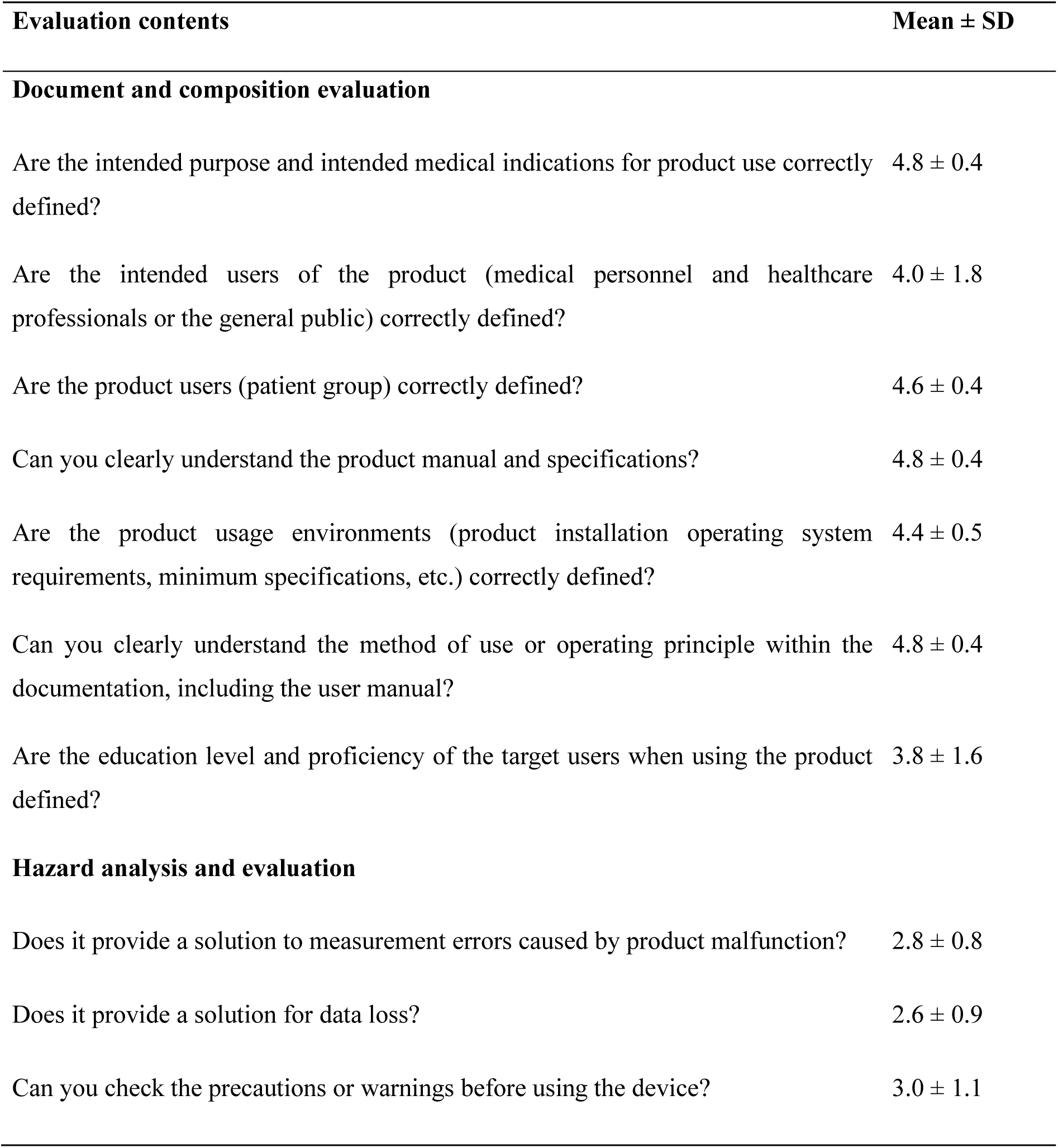
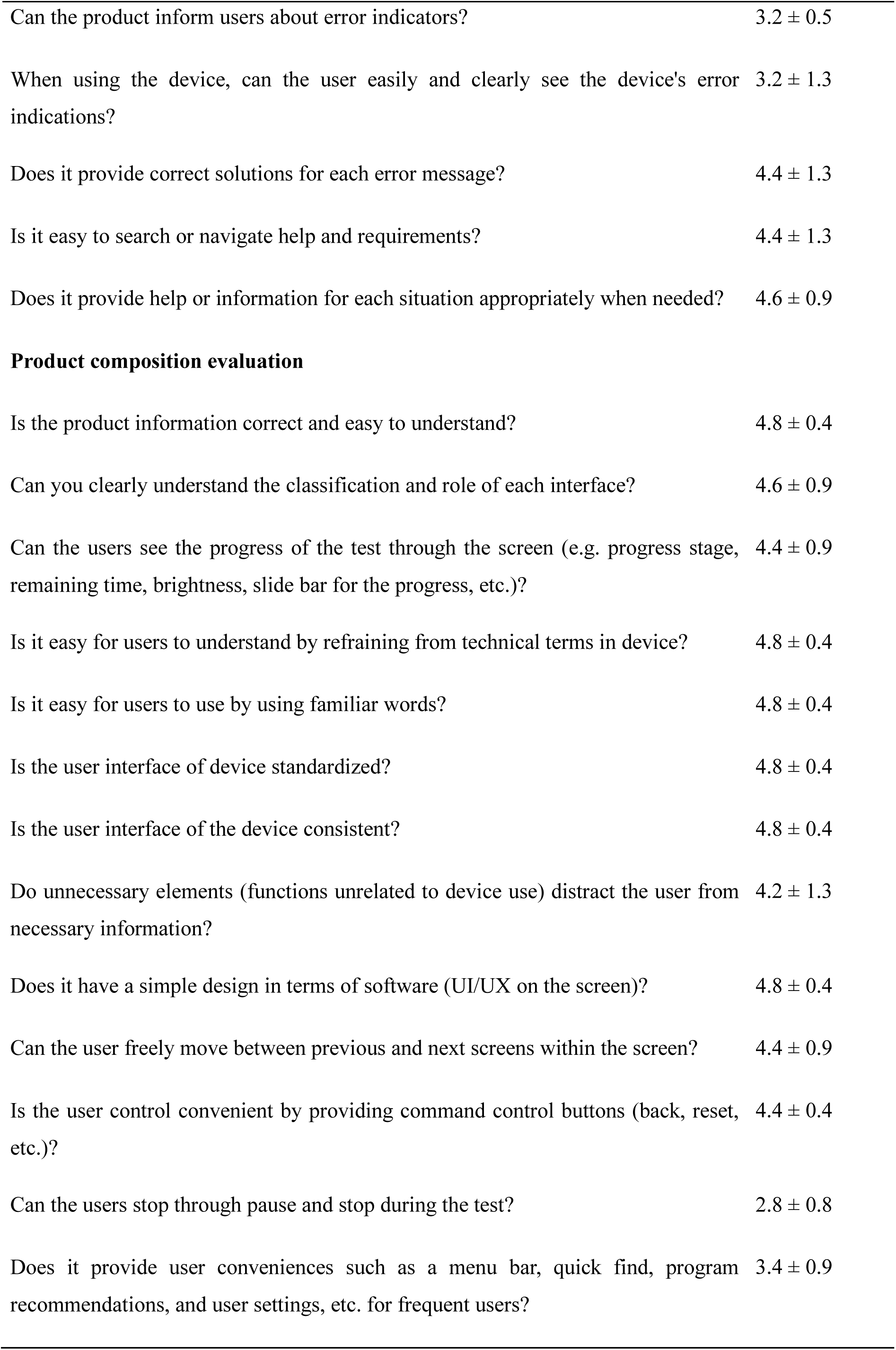
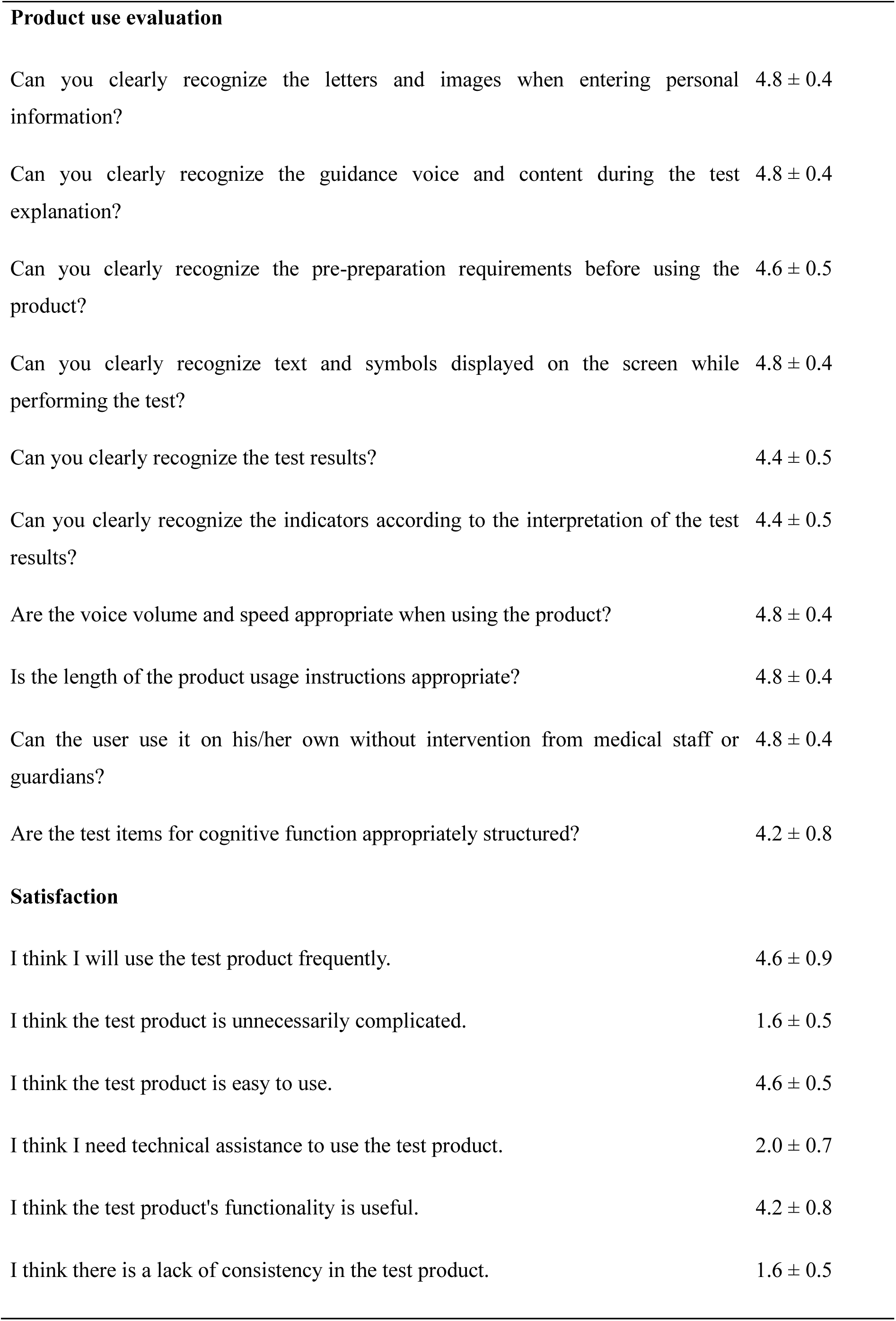
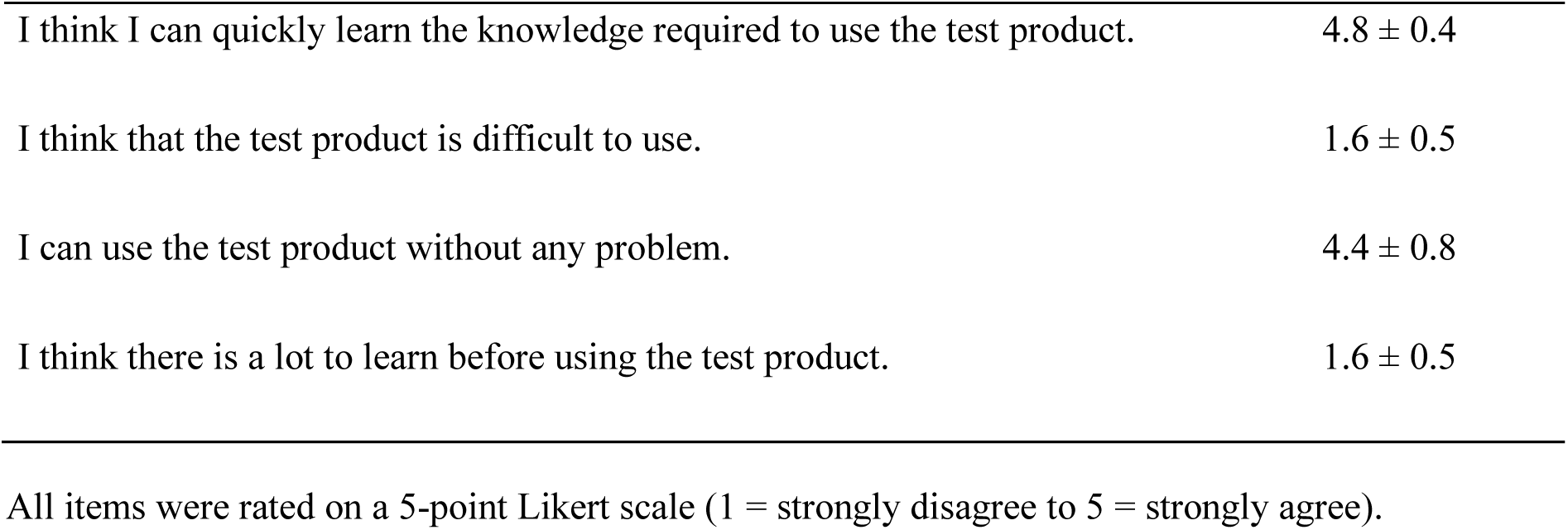
Results of the formative evaluation.

In the hazard analysis, the product demonstrated limited capability in addressing critical errors, such as solutions for malfunctions with 2.8 ± 0.8 and data loss with 2.6 ± 0.9. Moderate performance was observed in error notifications with 3.2 ± 0.5, while high scores were achieved in providing accessible help with 4.4 ± 1.3 and contextual information with 4.6 ± 0.9. These results suggest a need to refine error-handling mechanisms while leveraging the product’s strengths in user accessibility.

The Tool Composition evaluation revealed consistent strengths in interface design, with high scores for instructions at 4.8 ± 0.4, terminology at 4.8 ± 0.4, and interface consistency at 4.8 ± 0.4. Navigation between screens and accessibility of main buttons scored positively with 4.4 ± 0.9 and 4.4 ± 0.4, respectively. However, notifications during use were rated lower at 2.8 ± 0.8, highlighting an opportunity to improve user alerts.

The Usability Evaluation showed strong performance in clarity and accessibility, with personal information recognition scoring 4.8 ± 0.4, test instructions scoring 4.8 ± 0.4, and pre-test preparations scoring 4.6 ± 0.5. Test results and graphical representations scored slightly lower at 4.4 ± 0.5, suggesting enhancements to visual clarity. The cognitive test design was rated the lowest at 4.2 ± 0.8, indicating potential for refinement to better meet user expectations.

The Satisfaction Evaluation yielded a System Usability Scale score of 85.5, placing the product in the excellent usability category. Participants found the product easy to use with a score of 4.6, intuitive to learn with a score of 4.8, and cohesive in its functionality with a score of 4.2. Low scores for negative aspects, such as unnecessary complexity 1.6 and reliance on technical support with 2.0 reinforced its user-friendly design.

### Summative Evaluation

The summative evaluation (Table 3) demonstrated high usability and task completion across a ll scenarios. All participants, aged in their 50s and 60s, achieved a task success rate of 100% in most tasks, with only minor deviations observed in a small subset of tasks. Specifically, the personal information entry task showed a slightly lower success rate, reflecting occasional input errors. The System Usability Scale (SUS) scores ranged from 50 to 100, with an average score of 80.2, indicating strong overall usability. Errors, manual consultations, and help requests were minimal, emphasizing the program’s intuitive design.

**Table 3.**
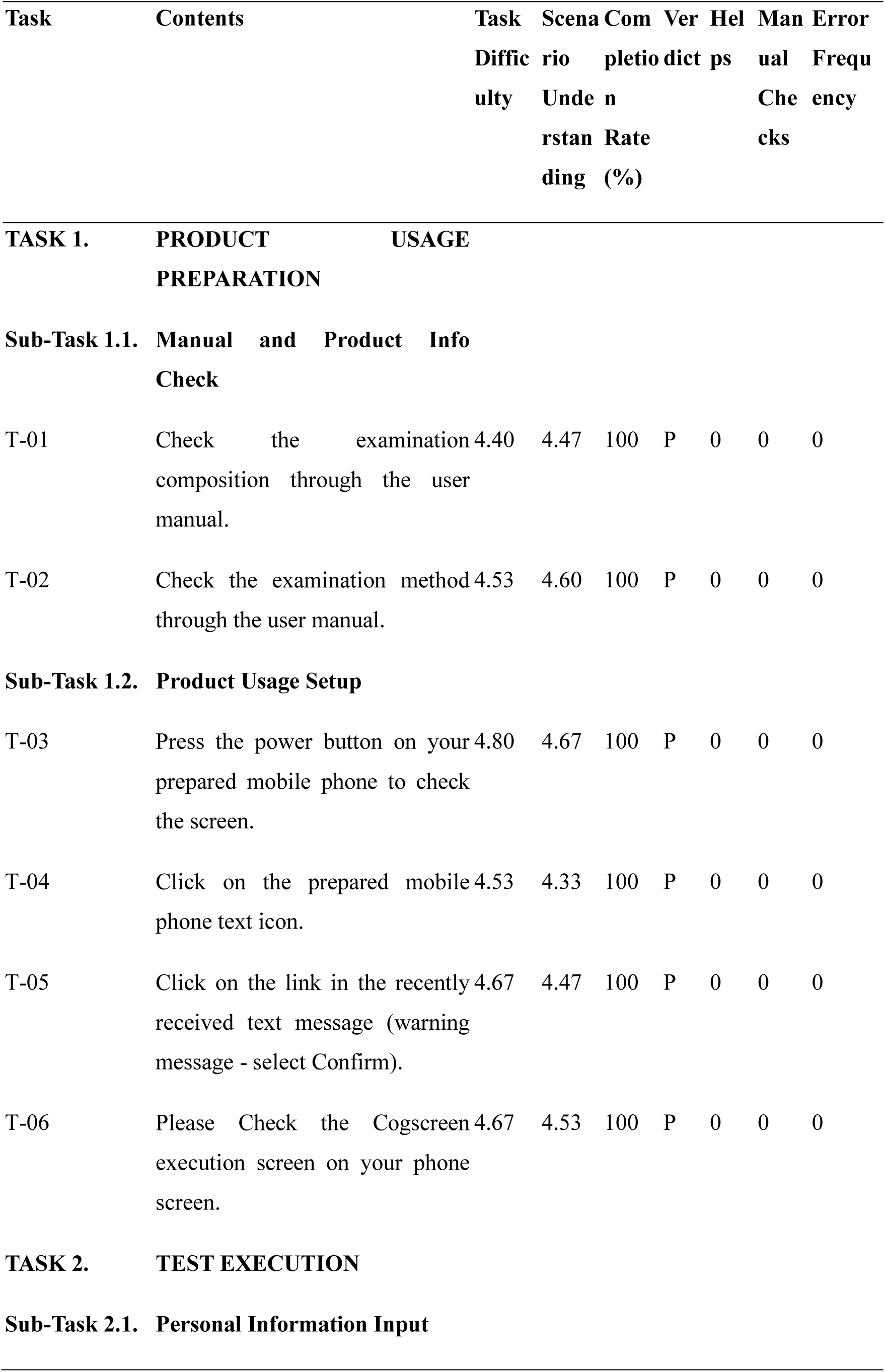
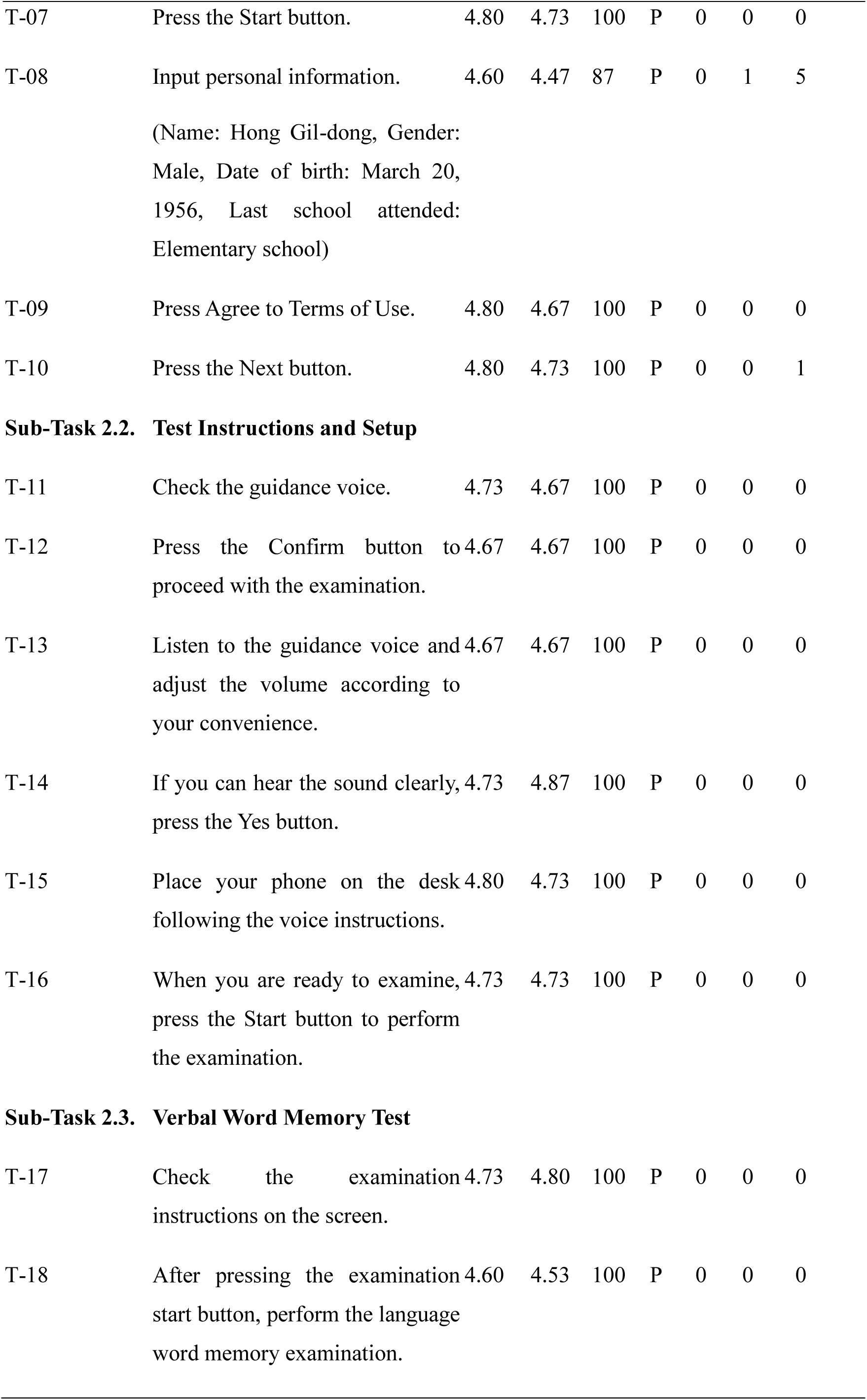
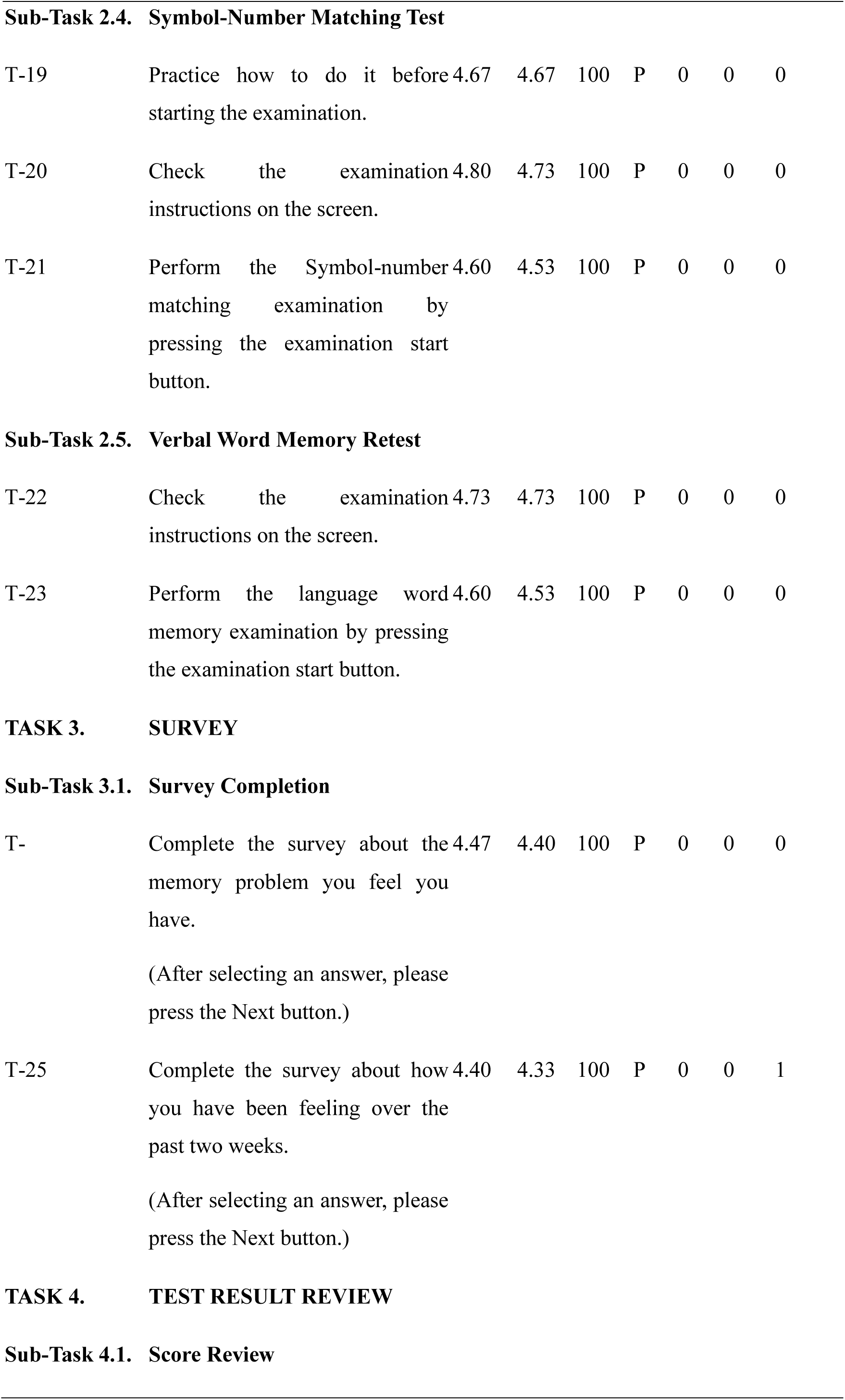
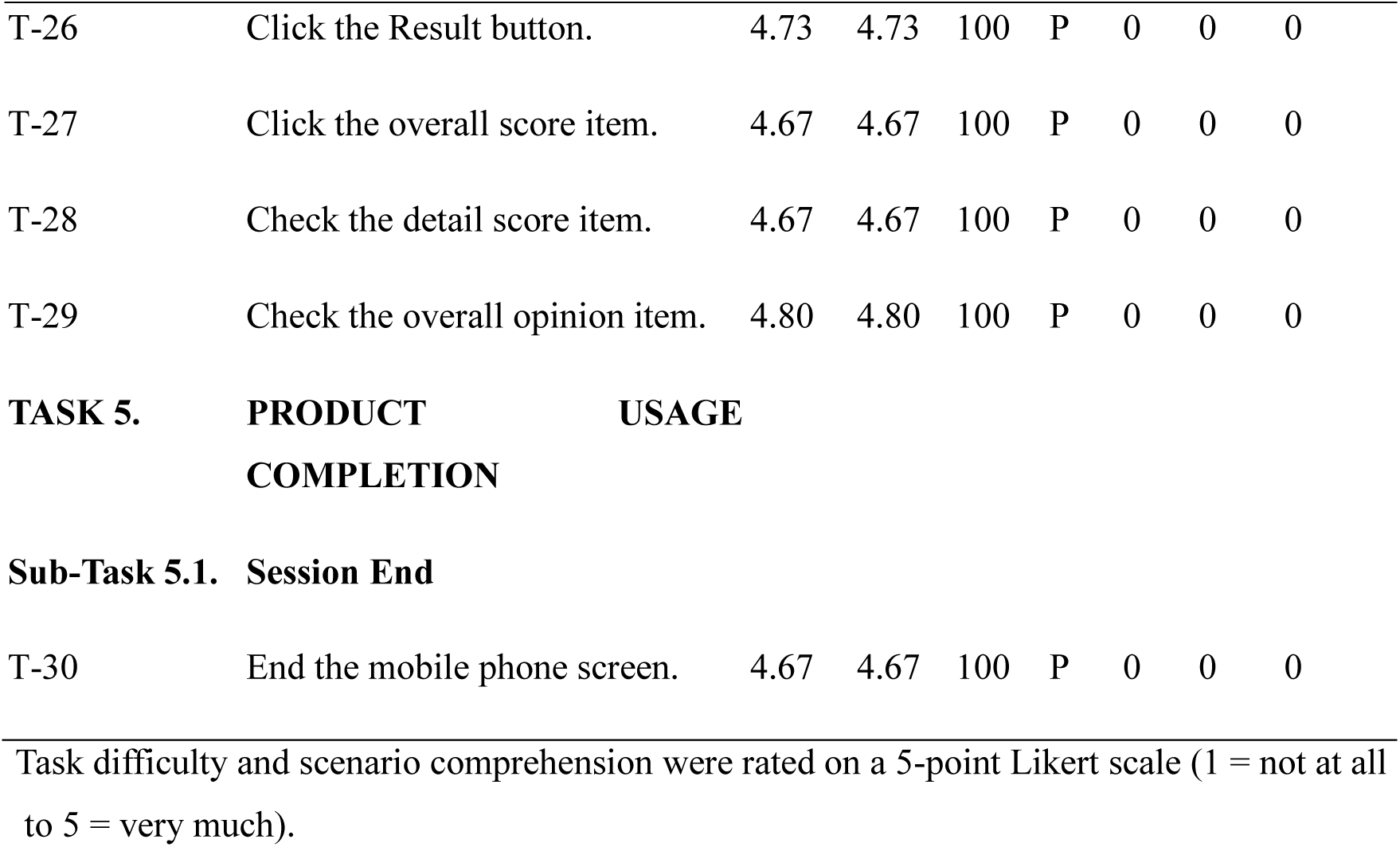
Task Scenario Outcomes and Usability for Cogscreen.

| Task | Contents | Task Difficulty | Scenario Understanding | Completion Rate (%) | Verdict | Helpfulness | Manual Checks | Error Frequency |
| --- | --- | --- | --- | --- | --- | --- | --- | --- |
| **TASK 1. PRODUCT USAGE PREPARATION** |  |  |  |  |  |  |  |  |
| **Sub-Task 1.1. Manual and Product Info Check** |  |  |  |  |  |  |  |  |
| T-01 | Check the examination composition through the user manual. | 4.40 | 4.47 | 100 | P | 0 | 0 | 0 |
| T-02 | Check the examination method through the user manual. | 4.53 | 4.60 | 100 | P | 0 | 0 | 0 |
| **Sub-Task 1.2. Product Usage Setup** |  |  |  |  |  |  |  |  |
| T-03 | Press the power button on your prepared mobile phone to check the screen. | 4.80 | 4.67 | 100 | P | 0 | 0 | 0 |
| T-04 | Click on the prepared mobile phone text icon. | 4.53 | 4.33 | 100 | P | 0 | 0 | 0 |
| T-05 | Click on the link in the recently received text message (warning message - select Confirm). | 4.67 | 4.47 | 100 | P | 0 | 0 | 0 |
| T-06 | Please Check the Cogscreen execution screen on your phone screen. | 4.67 | 4.53 | 100 | P | 0 | 0 | 0 |
| **TASK 2. TEST EXECUTION** |  |  |  |  |  |  |  |  |
| **Sub-Task 2.1. Personal Information Input** |  |  |  |  |  |  |  |  |
| T-07 | Press the Start button. | 4.80 | 4.73 | 100 | P | 0 | 0 | 0 |
| T-08 | Input personal information.<br>(Name: Hong Gil-dong, Gender: Male, Date of birth: March 20, 1956, Last school attended: Elementary school) | 4.60 | 4.47 | 87 | P | 0 | 1 | 5 |
| T-09 | Press Agree to Terms of Use. | 4.80 | 4.67 | 100 | P | 0 | 0 | 0 |
| T-10 | Press the Next button. | 4.80 | 4.73 | 100 | P | 0 | 0 | 1 |
| **Sub-Task 2.2. Test Instructions and Setup** |  |  |  |  |  |  |  |  |
| T-11 | Check the guidance voice. | 4.73 | 4.67 | 100 | P | 0 | 0 | 0 |
| T-12 | Press the Confirm button to proceed with the examination. | 4.67 | 4.67 | 100 | P | 0 | 0 | 0 |
| T-13 | Listen to the guidance voice and adjust the volume according to your convenience. | 4.67 | 4.67 | 100 | P | 0 | 0 | 0 |
| T-14 | If you can hear the sound clearly, press the Yes button. | 4.73 | 4.87 | 100 | P | 0 | 0 | 0 |
| T-15 | Place your phone on the desk following the voice instructions. | 4.80 | 4.73 | 100 | P | 0 | 0 | 0 |
| T-16 | When you are ready to examine, press the Start button to perform the examination. | 4.73 | 4.73 | 100 | P | 0 | 0 | 0 |
| **Sub-Task 2.3. Verbal Word Memory Test** |  |  |  |  |  |  |  |  |
| T-17 | Check the examination instructions on the screen. | 4.73 | 4.80 | 100 | P | 0 | 0 | 0 |
| T-18 | After pressing the examination start button, perform the language word memory examination. | 4.60 | 4.53 | 100 | P | 0 | 0 | 0 |
| **Sub-Task 2.4. Symbol-Number Matching Test** |  |  |  |  |  |  |  |  |
| T-19 | Practice how to do it before starting the examination. | 4.67 | 4.67 | 100 | P | 0 | 0 | 0 |
| T-20 | Check the examination instructions on the screen. | 4.80 | 4.73 | 100 | P | 0 | 0 | 0 |
| T-21 | Perform the Symbol-number matching examination by pressing the examination start button. | 4.60 | 4.53 | 100 | P | 0 | 0 | 0 |
| **Sub-Task 2.5. Verbal Word Memory Retest** |  |  |  |  |  |  |  |  |
| T-22 | Check the examination instructions on the screen. | 4.73 | 4.73 | 100 | P | 0 | 0 | 0 |
| T-23 | Perform the language word memory examination by pressing the examination start button. | 4.60 | 4.53 | 100 | P | 0 | 0 | 0 |
| **TASK 3. SURVEY****Sub-Task 3.1. Survey Completion** |  |  |  |  |  |  |  |  |
| T- | Complete the survey about the memory problem you feel you have.<br>(After selecting an answer, please press the Next button.) | 4.47 | 4.40 | 100 | P | 0 | 0 | 0 |
| T-25 | Complete the survey about how you have been feeling over the past two weeks.<br>(After selecting an answer, please press the Next button.) | 4.40 | 4.33 | 100 | P | 0 | 0 | 1 |
| **TASK 4. TEST RESULT REVIEW****Sub-Task 4.1. Score Review** |  |  |  |  |  |  |  |  |
| T-26 | Click the Result button. | 4.73 | 4.73 | 100 | P | 0 | 0 | 0 |
| T-27 | Click the overall score item. | 4.67 | 4.67 | 100 | P | 0 | 0 | 0 |
| T-28 | Check the detail score item. | 4.67 | 4.67 | 100 | P | 0 | 0 | 0 |
| T-29 | Check the overall opinion item. | 4.80 | 4.80 | 100 | P | 0 | 0 | 0 |
| **Sub-Task 5.1. Session End** |  |  |  |  |  |  |  |  |
| T-30 | End the mobile phone screen. | 4.67 | 4.67 | 100 | P | 0 | 0 | 0 |

Task difficulty and scenario comprehension were rated on a 5-point Likert scale (1 = not at all to 5 = very much).

Table 3 shows the results for task scenario outcomes and usability metrics. All tasks in the product usage preparation, test execution, survey completion, and result review phases achieved high completion rates, with scenario understanding scores consistently exceeding 4.5 on a Likert scale. Specific tasks, such as reviewing the user manual and performing memory tests, demonstrated high participant engagement and satisfaction. The structured design of tasks ensured minimal cognitive load, allowing users to navigate the system effectively. Qualitative feedback further supported these findings, with participants describing the system as accessible, intuitive, and valuable for cognitive assessment purposes. Recommendations for improvement focused on minor adjustments to enhance user convenience and accessibility.

Overall, the findings from Table 3 underscore the Cogscreen program’s robust usability in facilitating task completion. While minor refinements were suggested to optimize the user experience further, the high success rates and positive feedback indicate that the program is well-suited for its intended purpose. These results provide a strong foundation for future enhancements, ensuring the program remains accessible and effective for its target user population.

## Discussion

This study evaluated the usability and acceptability of Cogscreen, a digital screening tool developed to assess cognitive decline in elderly populations, particularly those with Mild Cognitive Impairment (MCI). By integrating objective cognitive tests and self-report scales of memory concerns and mood symptoms, Cogscreen aims to provide a holistic evaluation of cognitive health. Its mobile-based design facilitates independent use, making it a promising tool for addressing the accessibility barriers of traditional cognitive assessments. In contrast to tools focusing on single-domain tasks or requiring additional sensors, Cogscreen integrates both cognitive tasks and self-report measures of subjective decline and mood into a single mobile platform. Digital screeners such as CANTAB and Cogstate primarily emphasize task-based performance^9,10^, while DBFS demonstrates strong correlations with traditional paper-and-pencil tests but does not incorporate mood or subjective decline measures^11^. Other systems, including gait sensor-based or biomarker-driven platforms, require specialized hardware that may limit feasibility in primary care or community settings.^29,30^ Cogscreen differs in that it requires only a smartphone, offers a multidimensional assessment, and has been validated under international usability standards IEC 62366-1, FDA guidance.^27^ Nevertheless, these features do not fully eliminate common challenges such as digital literacy gaps or long-term adherence. Rather than replacing existing screeners, Cogscreen may be viewed as a complementary tool that prioritizes usability and independence while addressing some, but not all, of these recurring barriers.

The formative evaluation underscored Cogscreen’s clear purpose, intuitive design, and user-friendly documentation, indicating strong usability. At the same time, it highlighted areas requiring improvement, particularly support for users with lower digital literacy and the management of critical errors such as data loss and notification functions. These results demonstrate the importance of enhancing error management features to ensure robust usability. The summative evaluation confirmed Cogscreen’s feasibility as a self-administered screening tool. With nearly all tasks completed, participants were able to use the system independently. Minor difficulties, such as data entry and graphical inconsistencies, suggest refinements are needed to further enhance reliability. The SUS scores indicate its suitability for independent use.

Thus, within the landscape of digital screeners, Cogscreen occupies a complementary niche by combining accessibility and multidimensional assessment, while still requiring further validation in more diverse and impaired populations. By integrating objective and subjective measures, Cogscreen delivers a multidimensional assessment of cognitive and mood-related health, thereby offering distinct advantages over other instruments. A contribution of this study lies not only in demonstrating feasibility but also in applying both formative and summative usability evaluations in line with international standards IEC 62366-1, FDA guidance. Few digital cognitive tools have undergone such structured usability engineering, which strengthens the methodological novelty of this study. Beyond methodological novelty, the contribution of this study lies in combining objective cognitive tasks with self-report measures of subjective cognitive decline and mood. This multidimensional integration differentiates it from m any existing digital screeners that emphasize only task-based performance.

A limitation of this study is that the usability evaluation was conducted primarily with a relatively younger elderly population, which may not fully represent the usability challenges experienced by older or more cognitively impaired individuals. Future iterations should extend usability testing to include older adults, as well as MCI, to enhance generalizability and ensure broader applicability. Cogscreen’s potential integration with remote monitoring systems and healthcare platforms could further enhance its role in early detection and intervention for neurodegenerative diseases, solidifying its position as a critical resource in digital healthcare. Several limitations should be acknowledged. Although the number of participants (n = 15) was aligned with FDA human factors usability guidance^28^, no formal statistical sample size estimation or power analysis was conducted, and the findings should therefore be interpreted with caution. In addition, the study population was limited to cognitively normal older adults aged 50-65 years, which restricts the generalizability of the results to individuals with mild cognitive impairment or dementia. Furthermore, the mean and standard deviation for age could not be reported due to the categorical nature of the collected data. The evaluation was conducted over a short period, precluding the assessment of long term usability or learning effects. Moreover, the summative evaluation was carried out in a controlled clinical setting rather than in everyday practice environments, which may not fully capture real-world usage. Future studies with larger, clinically diverse cohorts, extended follow-up, and real-world deployment are warranted to validate the generalizability and long-term applicability of these findings.

Clinically, Cogscreen has the potential to serve as an accessible and scalable screening tool that could facilitate early identification of cognitive decline in community and primary care settings, where traditional assessments are often impractical. By enabling self-administered testing, it may help reduce the burden on healthcare professionals and increase opportunities for routine monitoring among at-risk populations. From a research perspective, the usability findings presented here establish a foundation for future validation studies in larger and more diverse cohorts, including individuals with MCI or dementia. Moreover, Cogscreen could be incorporated into longitudinal and intervention studies to track cognitive trajectories and evaluate the effects of preventive or therapeutic strategies in real-world environments.

While Cogscreen demonstrated strong overall usability, our findings highlight several challenges that are relevant to digital cognitive screening tools and should be addressed to ensure successful adoption. Strengthening error management through clearer notifications and safeguards against data loss remains essential to maintain user trust and data integrity. Streamlining data entry procedures and improving the presentation of results could reduce cognitive load and minimize errors, thereby enhancing overall efficiency. Moreover, providing tailored guidance and training resources for individuals with lower levels of digital literacy is essential for promoting equitable access and widespread use in diverse real-world populations.

As an initial step, this study demonstrates the overall usability of Cogscreen and provides important insights into the facilitators and barriers to its adoption, including its potential for independent use and challenges related to digital literacy and error management. These findings strengthen the evidence bas e supporting Cogscreen’s promise as a digital cognitive screening tool. However, further work is needed before widespread clinical deployment can be recommended. In particular, larger and more diverse clinical cohorts, evaluation in real-world practice settings, and refinements to the interface design will be essential to ensure successful implementation and sustainable integration into clinical workflows.

## Conclusion

The findings from both formative and summative evaluations highlight the strengths and areas for improvement in the design and functionality of Cogscreen. Overall results of both evaluations demonstrated that the tool effectively conveys its intended purpose and offers an elderly-friendly design, as reflected in high scores for clarity of instructions and intuitive interface. By addressing identified limitations and expanding usability testing to include more diverse populations, Cogscreen has the potential to become a critical resource for early detection and intervention in neurodegenerative diseases.

This study illustrates the essential role of iterative usability testing in the development of accessible, reliable, and effective digital health technologies tailored to the evolving needs of aging populations.

## Data Availability

All data produced in the present study are available upon reasonable request to the corresponding author.

## Acknowledgments

The authors thank all the participating patients and doctors.

## Author contributions/CRediT

Eunji Hwang led the drafting of the manuscript and compiled the evaluation results across both the formative and summative phases. Hyunsun Ham managed the overall evaluation process, including study design, participant coordination, and manuscript preparation. Dasom Lee contributed to the conceptual design of the Cogscreen content and supported the planning of the usability evaluations. Hanna Kim, Sang Hoon Oh, and Jun Gyu Kang contributed to the design and implementation of the formative and summative usability evaluations. Sehee Shim and Somin Lee developed the Cogscreen software application and supported methodological refinement. Jung-Hae Youn supervised the study framework and provided expert review of the evaluation protocols. Jun-Young Lee, as the corresponding author, provided strategic oversight throughout the development and evaluation process and contributed to the finalization of the manuscript.

## Ethical Approval

This study involved no patient data and posed minimal risk to participants. Institutional Review Board (IRB) review was therefore not required under the policies of Seoul National University College of Medicine. All participants provided written informed consent prior to participation. All participants provided written informed consent prior to participation.

## Guarantor

Jun-Young Lee is the guarantor of this work and accepts full responsibility for the integrity of the data and the accuracy of the data analysis.

