## Supplementary Materials for "User Experience Evaluation of Cogscreen for Screening Mild Cognitive Impairment: Formative and Summative Evaluation"

### Multimedia Appendix

**Table S1.** Results of the Summative Usability Evaluation

| Participant | Gender | Age Group | SUS SCORE | Task Success Rate | Number of Use Errors | Number of Manual Reference | Number of Help Request |
|---|---|---|---|---|---|---|---|
| P01 | Female | 60’s | 97.5 | 100 | 1 | 0 | 0 |
| P02 | Female | 60’s | 97.5 | 100 | 1 | 0 | 0 |
| P03 | Male | 50’s | 82.5 | 100 | 0 | 0 | 0 |
| P04 | Male | 60’s | 100 | 97 | 1 | 0 | 0 |
| P05 | Female | 50’s | 62.5 | 97 | 1 | 1 | 0 |
| P06 | Male | 60’s | 100 | 100 | 0 | 0 | 0 |
| P07 | Female | 60’s | 65 | 100 | 0 | 0 | 0 |
| P08 | Male | 60’s | 85 | 100 | 0 | 0 | 0 |
| P09 | Male | 60’s | 80 | 100 | 0 | 0 | 0 |
| P10 | Male | 60’s | 92.5 | 100 | 0 | 0 | 0 |
| P11 | Female | 60’s | 60 | 100 | 2 | 0 | 0 |
| P12 | Female | 60’s | 80 | 100 | 0 | 0 | 0 |
| P13 | Female | 60’s | 72.5 | 100 | 0 | 0 | 0 |

| | | | | | | | |
|---|---|---|---|---|---|---|---|
| P14 | Female | 60’s | 50 | 100 | 1 | 0 | 0 |
| P15 | Female | 50’s | 90 | 100 | 0 | 0 | 0 |

**Table S2**. Post-evaluation interview guide and participant responses after scenario implementation

| No. | Interview question | Participant | Opinion of the subject |
|---|---|---|---|
| 1 | Were the user manual clear enough and easy to understand? | P02 | It was easy. |
| | | P03 | It was very simple and easy. |
| | | P06 | It was easy to understand. |
| | | P07 | It was easy to understand. |
| 2 | What functions and elements would you like to see added when using the product? | P06 | I think I would remember more if I were informed in advance when taking a memory test. |
| | | P08 | It might be easier to go to “Power button” before starting and then “Enter personal information.” |
| | | P10 | It would be nice if a more precise description was added. (I think the first question in memory should specify finding memory.) |
| | | P15 | It would also be nice to have a system that could further enhance cognitive functions. |
| 3 | Have you experienced any discomfort or inconvenience while using this product? | | No opinion. |
| 4 | Is there anything you found convenient when using the product? | P01 | It’s easy. |
| | | P04 | I think it’s good because it can be done just by touching. |
| | | P07 | It is simple and clear, not complicated, so it is easy to use. |
| | | P10 | It’s convenient because you can just answer the questions as they are asked. |

| No. | Interview question | Participant | Opinion of the subject |
| --- | --- | --- | --- |
| | | P15 | I found it convenient because it was easy to solve simply. |
| 5 | Are there any areas where you feel there is inconvenience in using the product and need improvement? | P08 | I think there isn't enough time for "Subjective Cognitive Decline Questionnaire" or the "Depression Questionnaire." |
| | | P09 | I would like the answers to the questionnaire to be specific. |
| | | P15 | I think it would be good if the difficulty level were a bit higher. |
| 6 | Have you ever performed a similar digital cognitive examination on a mobile platform before? | P01–P15 | No. |
| 6-1 | If so, please tell me which product you used and where the examination was conducted. | | No opinion. |
| 6-2 | Please tell me what advantages or disadvantages this examination (Cogscreen) has compared to that product. | | No opinion. |
| 7 | Have you ever taken a similar cognitive examination on paper before? | P01–P15 | No. |
| 7-1 | If so, please tell me what examination you underwent and which institution conducted it. | | No opinion. |
| 7-2 | Please tell us what advantages or inconveniences this examination (Cogscreen) has compared to the paper examination. | | No opinion. |
| 8 | Free opinion | P01 | Since I am at an age where I need to be concerned about dementia and have a family history, I think it would be a good idea to use a product such as cognitive evaluation software. |
| | | P03 | It's my first time using it and it's convenient to use. |
| | | P04 | I feel like my cognitive abilities have declined a lot, so if I get the chance, I think it would be a good idea to |

| No. | Interview question | Participant | Opinion of the subject |
| --- | --- | --- | --- |
| | | | take a cognitive ability test. |
| | | P07 | This product is convenient and easy to use and seems to help improve memory and cognitive abilities in older adults. |
| | | P09 | I think it would be good to move the location of the answer bar (multiple choice) to the question up a little. |
| | | P11 | I wish the symbol test was a little clearer. |
| | | P15 | I look forward to a more specific and challenging evaluation. (It would be nice if there were high, medium, and low difficulty levels.) |
